# Ocular motor biomarkers in Niemann-Pick disease type C: A prospective cross-sectional multicontinental study in 72 patients

**DOI:** 10.1101/2020.08.05.20158915

**Authors:** Tatiana Bremova-Ertl, Larry Abel, Mark Walterfang, Ettore Salsano, Anna Ardissone, Věra Malinová, Miriam Kolníková, Jordi Gascón Bayarri, Ali Reza Tavasoli, Mahmoud Reza Ashrafi, Yasmina Amraoui, Eugen Mengel, Stefan A. Kolb, Andreas Brecht, Stanislavs Bardins, Michael Strupp

**Affiliations:** Department of Neurology, University Hospital Bern (Inselspital) and University of Bern, Bern, Switzerland; Center for Rare Diseases, Institute for Clinical Chemistry, University Hospital Bern (Inselspital) and University of Bern, Bern, Switzerland; Optometry & Vision Science, School of Medicine, Deakin University, Waurn Ponds, VIC, Australia; Neuropsychiatry Unit, Royal Melbourne Hospital, Parkville, VIC, and; Melbourne Neuropsychiatry Centre, University of Melbourne & NorthWestern Mental Health, Parkville, VIC, Australia; Unit of Rare Neurodegenerative and Neurometabolic Diseases, Fondazione IRCCS Istituto Neurologico “Carlo Besta”, Milan, Italy; Department of Pediatrics and Adolescence Medicine, First Faculty of Medicine, Charles University, General University Hospital Prague, Czech Republic; Department of Child Neurology, Comenius University Children’s Hospital, Bratislava, Slovak Republic; Department of Neurology, Hospital Universitari de Bellvitge, L’Hospitalet de Llobregat, Spain; Myelin Disorders Clinic, Pediatric Neurology Division, Children’s Medical Center, Pediatric Center of Excellence, Tehran University of Medical Sciences, Tehran, Iran; Villa Metabolica, Center for Paediatric and Adolescent Medicine, University Medical Center of the Johannes Gutenberg University Mainz, Germany; SphinCS Gmbh, Clinical Science for LSD, Hochheim, Germany; Actelion, a Janssen company of Johnson & Johnsons, Switzerland; German Center for Vertigo and Balance Disorders and Department of Neurology, University Hospital Munich, Campus Grosshadern, Ludwig-Maximilians-University Munich, Germany

**Author notes:** **Corresponding author:** Tatiana Bremova-Ertl, MD, PhD, Department of Neurology, University Hospital Bern (Inselspital) and University of Bern, Bern, Switzerland, and. Statistical analysis was performed by Syntax for Science, Parc BIT, Ctra. Valldemossa, 07121 Palma de Mallorca, Spain. **Email addresses of co-authors:**.

**Keywords:** Niemann-Pick Type C, ocular motor function, saccades, biomarkers, supranuclear vertical saccade palsy, supranuclear vertical gaze palsy, video-oculography

## Abstract

Niemann-Pick type C (NPC) is a rare lysosomal storage disorder with ocular motor involvement. In a multicontinental, cross-sectional study we characterized ocular motor function in 72 genetically proven patients from twelve countries by means of video-oculography. Interlinking with disease severity, we also searched for ocular motor biomarkers. Our study protocol comprised reflexive and self-paced saccades, smooth pursuit, and gaze-holding in horizontal and vertical planes. Data were compared with those of 158 healthy controls. The Modified Disability Rating Scale, Scale for Assessment and Rating of Ataxia, Spinocerebellar Ataxia Functional Index for neurological status, and Montreal Cognitive Assessment for cognition were also performed.

In contrast to previous publications and the common belief that the “downward saccadic system degenerates to greater extent than the upward one”, our measurements of vertical saccades demonstrated that the involvement in both directions was similar. Mean saccadic peak velocity to 20° stimulus was 63.5°/s (SD, 95% CIs of the mean: 59.5, [47.9-79.2]) in NPC patients and 403.1°/s (69.0, [392.0-414.2°/s]) in healthy subjects (p<0.001). Downward saccades yielded 51°/s (68.9, [32.7-69.3]), whilst upward 78.8°/s (65.9, [60.8-96.8]) (p<0.001). Vertical position smooth pursuit gain was 0.649 (0.33, [0.554-0.744]) in NPC and 0.935 (0.149 [0.91-0.959]) in HC (p<0.001).

The number of patient-specific saccadic patterns, incl. slow-pursuit like, hypometric and staircase-pattern saccades suggest varying involvement of the saccadic system with fragmentation of the velocity profile as a sign of omnipause neuron dysfunction. Observed compensating strategies, such as blinks to elicit saccades, head and upper body movements to overcome the gaze palsy, should be used clinically to establish a diagnosis.

Vertical reflexive saccades were more impaired and slower than self-paced ones. Ocular motor performance depended on age of onset and disease duration.

We found that peak velocity and latency of horizontal saccades, vertical saccadic duration and amplitude, and horizontal position smooth pursuit can be used as surrogate parameters for clinical trials, as they showed the strongest correlation to disease severity. By comparing saccadic with pursuit movements, we showed that 98.2% of patients generated vertical saccades (both up and down) that were below the 95% confidence intervals of the controls’ peak velocity. Only 46.9% of patients had smooth pursuit gain lower than that of 95% of healthy controls. Vertical supranuclear *saccade* palsy and not vertical supranuclear *gaze* palsy is the hallmark of NPC disease. The distinction between saccadic and gaze palsy is also important in other neurodegenerative diseases and inborn errors of metabolism with ocular motor involvement, such as progressive supranuclear palsy or Gaucher disease type 3.

## Introduction

Eye movement abnormalities are sensitive markers in a variety of neurodegenerative disorders. One such rare, autosomal recessive inborn error of lipid metabolism is Niemann-Pick type C (NPC), caused by a mutation in the NPC1 (95%) or NPC2 genes. Defects result in a cellular accumulation of endocytosed unesterified cholesterol and glyco- and sphingolipids in the late endosome/lysosome, leading to functional disturbance of neural and non-neural tissues (Vanier, 2015). Characteristic of NPC is the heterogeneity of manifestations that comprise visceral, neurological and psychiatric signs. Often symptoms are not disease specific, arise at different ages, and progress at different rates (Patterson *et al*., 2017).

In NPC, it is known that vertical eye movements are affected much earlier in the course of the disease than horizontal eye movements (Abel *et al*., 2009, 2012). In the past, vertical supranuclear *gaze* palsy (VSGP), but not vertical supranuclear *saccade* palsy (VSSP) was highlighted as the hallmark symptom of NPC at all disease stages, not recognizing the actual ocular motor impairment, even though the predominant vertical *saccadic* impairment was described in several case-studies (Rottach *et al*., 1997; Solomon *et al*., 2005; Salsano *et al*., 2012). Vertical supranuclear *saccade* palsy is caused by the impairment of burst neurons in the rostral interstitial nucleus of the medial longitudinal fascicle (riMLF) in the mesencephalon (Horn and Buttner-Ennever, 1998). The burst neurons in the riMLF play a key role in the generation of vertical saccades and vertical quick phases of nystagmus. Vertical supranuclear *gaze* palsy denotes a combined dysfunction of both saccades *and* smooth pursuit. It is caused by an additional impairment of the Interstitial Nucleus of Cajal (InC) in the rostral midbrain and its connections through the posterior commissure (Büttner-Ennever and Horn, 2002; Büttner-Ennever *et al*., 2003).

For the approval of miglustat for the treatment of NPC, an impairment of horizontal saccadic function was used as a surrogate marker (Patterson *et al*., 2007). Until now, however, only single cases or small cohorts of NPC patients were thoroughly examined in terms of saccadic function (Rottach *et al*., 1997; Solomon *et al*., 2005; Abel *et al*., 2009, 2012, 2015) and little is known about the remaining ocular motor systems and their deficits in terms of disease severity. In contrast to impaired eye movements, vestibular function is intact in patients with NPC (Bremova *et al*., 2016). The ocular motor abnormalities correlate to the neurodegenerative changes in the retina (Havla *et al*., 2020).

This was a cross-sectional prospective study in a large international cohort of patients with NPC at different stages of the disease with diverse mutation backgrounds to account for possible genetic impact. Our objectives were as follows:

*First*, to comprehensively characterize ocular motor function on cerebral, cerebellar and brainstem levels. This was achieved by examining reflexive and self-paced saccades, smooth pursuit and gaze holding in vertical and horizontal plane, respectively.

*Second*, to clarify whether VSSP or VSGP is the hallmark of NPC disease. This is of importance in clinical practice. Due to the limited amount of time per patient, clinicians perform specific examinations, often checking for the presence of VSGP, instead of VSSP, leading to a considerable diagnosis delay.

*Third*, to correlate ocular motor dysfunction with the severity of disease to identify surrogate biomarkers that track the progression of the disease and quantify the therapeutic benefits in clinical trials. Most importantly, we built up univariant linear regression models to predict disease progression based on the relevant measures and to account for heterogeneous disease characteristics.

## Materials and methods

### Patients and control subjects

Out of 82 patients screened, 72 patients with genetically and/or biochemically confirmed NPC disease (30 Female, 42 Male, age ± SD 28.7 ± 14.2 years, mean disease duration 13.2 ± 9.0 years, mean age at the establishing of the diagnosis 22.2 ± 15.3 years) were included. These patients were originally (not referring to study sites) from Germany (32.46%), Slovakia (1.44%/2 patients), Czech Republic (15.3%), Greece (2.8%), Italy (5.6%), Bulgaria (0.72%/1 patient), Saudi Arabia (0.72%/1 patient), Spain (18%), Sweden (0.72%/1 patient), Turkey (1.44%/2 siblings), Iran (12.5%), and Australia (8.3%). The characteristics of all patients are listed in the supplemental table (**Supplemental Table S1**). The mean age at first symptoms of the disease was 14.7 years (SD ± 95% CI 11.1, [12.1-17.3]) **(Fig. 1)**. The first neurological symptoms appeared at the age of 16.6 years (12.9, [13.5-19.7]) and the first psychiatric symptoms appeared at the age of 19.0 years (10.4, [14.9-23.1]). The BMI of patients based on age of onset was 16.6 kg/m^2^ in the early-infantile form (n = 2). In the late-infantile form of the disease, the mean BMI was 16.1 ± 6.2 (n = 4), 21.5 ± 5.1 (n = 26) in the juvenile form and 23.7 ± 3.7 (n = 31) in the adult form.

**Figure 1.**
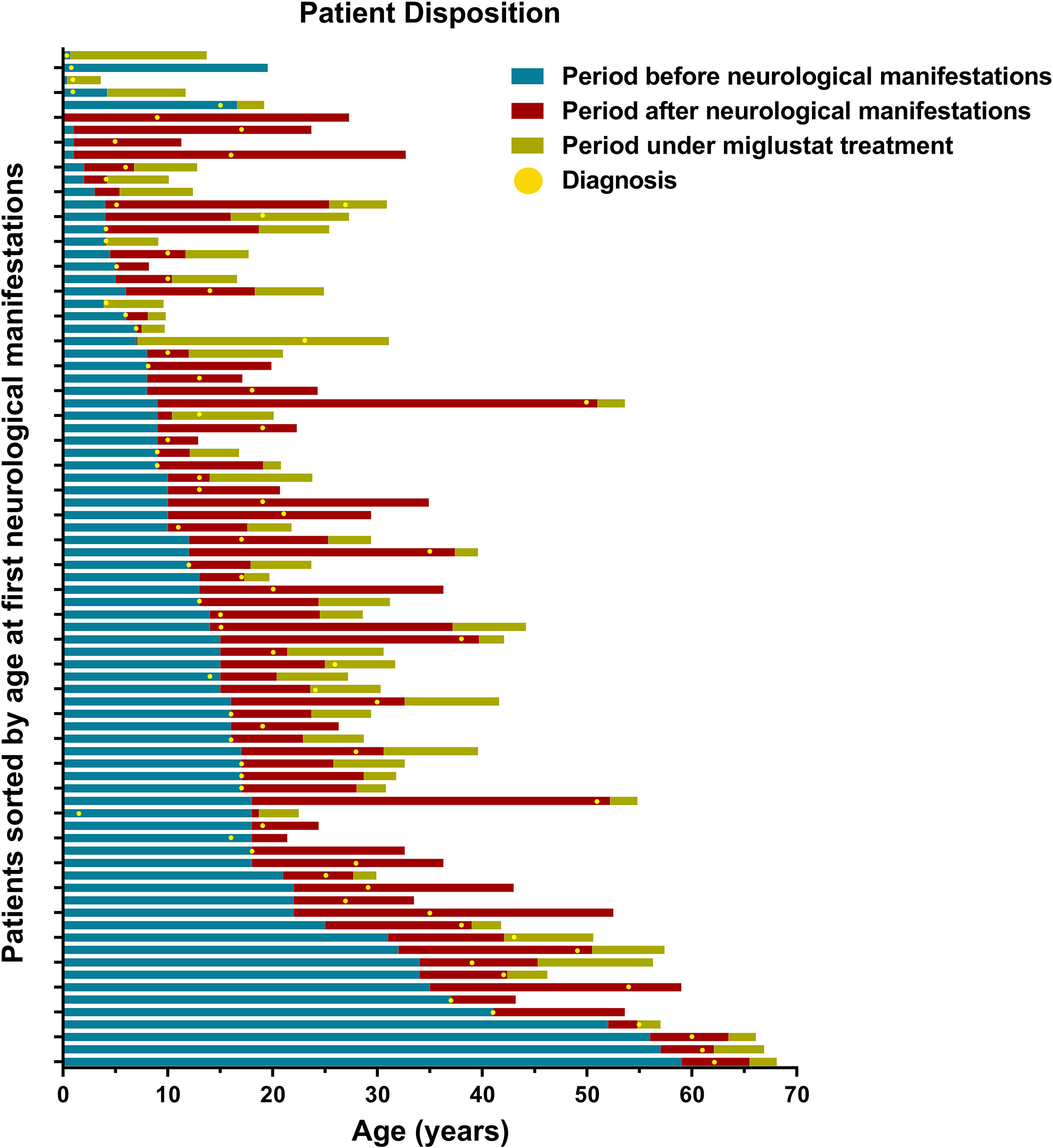
Patient disposition based on age at first neurological manifestation Fig 1. Overview of patient and disease characteristics based on the age at first neurological sign (ascending). Characteristics of all 82-screened patients are included.

Sixty-eight out of the 72 included patients had a mutation in NPC1, out of which a single patient had an additional mutation in NPC2 gene p.E20* **(Supplemental Table S1)**. Seventy-two patients were compliant with the examination; the compliance of the remaining patients was restricted because of cognitive impairment or disease severity. In the 10 patients without analyzable VOG results, the ocular motor deficits with compensating strategies (blinks, head/upper body thrusts) were profound, generating ocular motor data of insufficient quality (noisy data). These patients were excluded from the final analysis. For mutation background, see **Supplemental Table S1**. The highly frequent mutations p.I1061T and p.P1007A were present just in one patient (p.I1061T). Filipin staining was performed in 64.6 % of all patients: it was variant in 20% and classical in 30.8%. In addition, to establish the diagnosis and disease development, elevated levels of lysosphingolipids were detected in 9 (9.7%), oxysterols in 17 (23.6%), and, most commonly, chitotriosidase in 25 patients (34.7%). Most patients were on medication with miglustat (62 patients, 86.1%). The mean duration (±SD, 95% CI) of miglustat treatment was 3.9 years (3.1, [2.9-5.0]). Twenty-eight (68.3%) were on miglustat for longer than one year, and 13 (31.7%) for less than one year.

Regarding the NPC subtype, 2 patients (3.2%) were early-infantile, 5 (7.8%) late-infantile, 26 (40.6%) juvenile, and 39 patients (48.4 %) adult (>15 years of age). Twenty-four patients (34.3 %) had a first degree relative with NPC disease.

Since the disease manifests with a wide range of neurological symptoms **(Table 1)**, patient medication often included a number of CNS-active pharmaceutical agents. These included antiepileptics (9 patients, 12.5%), antidepressants (11, 15.3%), nootropics (7 patients, 9.7%), antipsychotics (10 patients, 13.9%), anti-Parkinsonian medication (including anticholinergics) and benzodiazepines (4 patients, 5.6%). Three patients had hearing devices due to hearing loss. Acetyl-DL-leucine was administered in twenty-one patients (31.7%) for cerebellar ataxia (Bremova *et al*., 2015). Two of these patients received this treatment for more than one year. One patient was treated with cyclodextrin, while ursodeoxycholic acid (UDCA) was administered to a further patient (early-infantile case with profound visceral manifestation).

**Table 1.** Clinical neurological and psychiatric manifestations in Niemann-Pick type C patients. Percentage of neurological and psychiatric manifestations in patients with Niemann-Pick type C (NPC). Note the percentage of clinically noted vertical supranuclear gaze palsy (VSGP) and/or vertical supranuclear saccade palsy (VSSP) that is much lower than detected video-oculographically.

| NPC patients |  |
| --- | --- |
| Patients with neurological manifestations |  |
| Vertical Supranuclear Gaze Palsy (VSGP), including Vertical Supranuclear Saccade Palsy (VSSP) | 41 (56.9%),<br>48 (59.3%) |
| Gelastic cataplexy | 9 (12.7%) |
| Clumsiness or frequent falls | 52 (72.2%) |
| Dysdiadochokinesis | 49 (70.0%) |
| Dysmetria | 50 (71.4%) |
| Intentional tremor | 29 (42.0%) |
| Dysarthria and/or dysphagia | 63 (88.7%) |
| Dyskinesia | 26 (37.7%) |
| Dystonia | 48 (67.6%) |
| Acquired and progressive spasticity | 15 (21.1%) |
| Hypotonia | 9 (12.7%) |
| Delayed developmental milestones | 13 (18.3%) |
| Seizures (partial or generalised) | 15 (20.8%) |
| Myoclonus | 9 (12.5%) |
| IQ (n = 16) | 64.8 (17.8, [55.3, 74.2])<br>Mean (SD, CI) |
| Patients with psychiatric manifestations |  |
| Cognitive decline in children or dementia in adults | 62 (86.1%) |
| Psychotic symptoms (hallucinations, delusions and/or thought disorder) | 16 (19.5%) |
| Treatment-resistant psychiatric symptoms | 9 (12.5%) |
| Disruptive or aggressive behavior | 24 (29.3%) |
| Progressive psychiatric disorders | 11 (15.3%) |
| Other psychiatric disorders | 14 (19.7%) |

Data collection was similar to that of International NPC Registry with demographic information (**Figure 1, Supplemental Table S1**). We performed an observed case analysis, with all summary and percentages calculated relative to the number of patients with available ocular motor data. In the resulting statistics, the denominators for analysis were the numbers of patients with the corresponding data.

One hundred and fifty-eight control subjects with no presence of neurological, ophthalmological or vestibular disease (55 F, age ± SD 41.7 ± 24.3 years) were also included to build up 95% confidence limits using the same testing paradigms.

The study was performed in accordance with the Helsinki II Declaration and approved by the ethics committee of the Ludwig Maximilian University Medical Faculty (addendum to project no 379-12). All participants or their legal guardians gave their informed consent prior to their inclusion in the study.

### Eye movement recordings

Stimuli were viewed binocularly in a dimly lit room, and the two-dimensional movements of the left eye were recorded, given conjugate gaze in NPC patients. Eye dominance was not determined. Eye movements were recorded using a video-based eye-tracker system (EyeSeeCam®, Munich, Germany). Head movements were also recorded. Algorithms for the calculation of eye position were based on the determination of the center of the pupil (ellipse fit). Eye movement recordings were sampled at intervals of 4.5 ms (220 Hz) with spatial resolution of approximately 1 minute of arc depending on pupil size, which by an image size of 188*120 pixels yielded a noise level of 0.015° (Schneider *et al*., 2009). The instrument was calibrated for each participant by recording fixations at the central and eccentric positions across a range of ±8.5° array horizontally and vertically. The participant’s head was steadied on the chinrest with the temporal bones restrained on both sides. The chinrest was adjusted so that the eyes were in the primary position when looking at the center of the calibration array. Although refractive errors were not corrected during data collection, it was verified at the time of calibration that all participants were able to see the targets well enough to localize them without any difficulty. The visual targets were displayed on a 22” computer monitor (LG, FLATRON W2242PK-SS, LG Electronics, Germany) running at 60 Hz. Monitor luminance was 250 cd/m^2^. The monitor was 60 cm from the bridge of the participant’s nose and subtended a visual angle of 43.2° horizontally by 27.7° vertically.

### Ocular motor paradigms

The following paradigms were performed in the same chronological sequence in all subjects, beginning with vertical, followed by horizontal eye movement recordings: reflexive and self-paced saccades, smooth pursuit, and gaze-holding.

Before each test, task instructions were given by the investigator and subjects were asked whether they understood the instructions. If the subject did not understand the instructions, they were repeated and demonstration trials were shown.

*Saccades:* All subjects showed visually guided vertical and horizontal pro-saccades in response to stimuli of 1.33° visual angle (expression changing smileys). Vertical saccades, elicited by the stimuli of 10° and 20° amplitude over the range ±10° from the central position and horizontal saccades of 5°, 15° and 30° amplitude, over the range ±15° from the central position, were required. Participants performed seven saccades in response to the stimulus of each size along both axes. The targets were presented in pseudorandom order for the time of 2500 ms with additional variation of 500 ms. Patients were provided with verbal encouragement to follow the target jumps and, for patients with more advanced disease, the investigator, or administrating care giver, pointed to target locations.

*Self-paced saccades:* We instructed subjects to switch arbitrarily between presented visual stimuli as quickly as possible (“make it a race”). Visual stimulation was permanently visible with two dots at 10° up and down, with stimulus amplitude of 20° in the vertical plane. Horizontally, there were two permanently visible dots at 15° on the right and left side, with a stimulus amplitude of 30°. Test duration was 30 seconds. All the parameters of reflexive saccades except the intersaccadic interval (not latency *per se*, since responses are internally triggered rather than being externally cued) and the absolute number of saccades (analyzed as a number of detected saccades after exclusion of blinking artefacts) were analyzed (for parameters, s. section Data Analysis, Ocular motor data).

*Smooth pursuit:* After the initial fixation period of 2 s, the target subtending visual angle of 0.57° moved in 3 cycles at 0.1 Hz and 3 cycles at 0.2 Hz frequencies, yielding peak target velocities of 9.5°/s, 18.8°/s horizontal and 6.4°/s, 12.6°/s vertical. The amplitude horizontally (right and left) comprised ±15° from the central position, and then ±10° vertically (up and down) without a break.

*Gaze-holding:* Participants were asked to foveate a point of 0.57° visual angle presented on a monitor, without moving their head. The point was primarily positioned at the level of the eyes, then changed the eccentricity from 15° left to 15° right and 10° down and 10° up from the central point, respectively. The stimulus was presented for 10 s at each position.

### Clinical evaluation

For the clinical characteristics of individual patients, see **Supplemental Table S1**.

#### Neurological examination

To assess the disease severity, the modified Disability Rating Scale (mDRS) was applied (Iturriaga *et al*., 2006). MDRS is a 4-domain scale (ambulation, manipulation, language, and swallowing) in an extended form (Pineda *et al*., 2010), which also includes seizures and ocular movements.

Cerebellar function was evaluated by administering the Scale for the Assessment and Rating of Ataxia (SARA) (Subramony, 2007; Weyer *et al*., 2007), an eight-item clinical rating scale (gait, stance, sitting, speech, fine motor function and taxis; range 0-40, where 0 is the best neurological status and 40 the worst). Moreover, patients were assessed by the Spinocerebellar Ataxia Functional Index (SCAFI), comprising the 8-Meters-Walking-Test (8MW), performed by having patients walking as quickly as possible from one line to another excluding turning twice, 9-Hole-Peg-Test (9HPT) of both dominant (9HPTD) and non-dominant hand (9HPTND) and the number of “PATA” repetitions over 10 s (PATA) (Schmitz-Hübsch *et al*., 2008).

#### Neuropsychological examination

To evaluate cognitive function, the Montreal Cognitive Assessment (MoCA) (Nasreddine *et al*., 2005) was used, which assesses different cognitive domains, including attention and concentration, executive functions, memory, language, visuoconstructional skills, conceptual thinking, calculations, and orientation. This tests has a total of 30 points and a cut-off score of 26 points.

#### Exploratory genotype-ocular motor phenotype correlation

Since the NPC1 gene has 3834 nucleotides (1278 amino acids) and there are mutations at 101 different positions (and multiple combinations in case of compound heterozygotes), seven categories were created based on the respective NPC protein domain. The limitation of this analysis was that compound heterozygotes (85.2% of all patients; in seven patients, the mutations were unknown or not indicated) were automatically assigned to two groups, thus generating a false total number of 98 cases. We took this aspect into account by creating the additional categories. The analyzed categories and their respective patient representation (n = 72, n (%)) were as follows (Allele 1/Allele 2): Cholesterol Binding (NTD) 2 (2.8%), Cholesterol Binding (NTD)/Cysteine-rich domain (CTD) 2 (2.8), Cysteine-rich domain (CTD) 12 (16.7%), Loop 2 (Middle Luminal Domain) 5 (6.9%), Loop 2 (Middle Luminal Domain)/Cysteine-rich domain (CTD) 10 (13.9%), Steroid Sensing Domain Transmembrane Helices (TM3-8) 6 (8.3%), Steroid Sensing Domain (TM3-8)/Cysteine-rich domain (CTD) 4 (5.6%), Steroid Sensing Domain (TM3-8)/Loop 2 (Middle Luminal Domain) 2 (2.8%), TM1-TM2 & Cysteine-rich domain (CTD) 1 (1.4%), TM1-TM2 & Loop 2 (Middle Luminal Domain) 1 (1.4%), TM1-TM2 & Steroid Sensing Domain (TM3-8) 1 (1.4%), TM9-TM13+COOH 6 (8.3%), TM9-TM13+COOH/Cysteine-rich domain (CTD) 12 (16.7), TM9-TM13+COOH/Loop 2 (Middle Luminal Domain) 2 (2.8%), and Unknown/Not-indicated 6 (8.3%).

## Data availability

The datasets used and analyzed during the presented study are available from the corresponding author upon reasonable request.

## Data analysis

Data were stored for off-line analysis and analyzed with MATLAB (The Mathworks Inc.), using the signal processing, image processing, and statistics toolboxes.

### Ocular motor data

*Saccades:* Saccades were marked automatically with a velocity threshold technique and verified manually for artifacts and accuracy. A saccade was considered to have been made in response to a specific stimulus, if the following criteria were met: 1) The saccade was the first saccade after the stimulus (first extrema selected), 2) it was in the correct direction, 3) the saccade velocity was higher than 5°/s (velocity threshold) between target presentation and saccade onset and 4) the saccade onset and offset was defined with a velocity of 2°/s. The program found these data points automatically. In order to reliably exclude saccades with blinks and artifacts, every saccade was visually evaluated and, if necessary, the saccade onset/offset was manually determined.

The following saccadic parameters were analyzed: 1) *Peak Velocity (PV)*, defined as maximal velocity during the saccade. To study the velocity of saccades, the “main sequence” plots (Bahill *et al*., 1975) of peak velocity versus amplitude and duration were built (Garbutt *et al*., 2003), as applied previously in other neurodegenerative disorders (Chen *et al*., 2010). 2) *Latency*, defined as the interval between target presentation and when the eye velocity reached 5°/s. *3) Intersaccadic interval*, defined as time difference between onset of the first and onset of the subsequential saccade. 4) *Amplitude*, defined by the position of the eye at the start of the saccade and the position of the final gaze shift. 5) *Gain*, defined as a ratio of the saccade amplitude to stimulus amplitude. 6) *Duration of the saccade*, time between the onset and the end of saccade. 7) *Peak duration*, defined as time to reach the peak velocity. Saccades in response to 5° were too small to be clearly recognized from the background noise generated by patients’ compensating strategies, so they were excluded from the final analysis.

Linear regression lines were fitted to the data of saccade peak duration versus amplitude to build up a *Slope of Peak Duration vs Amplitude*.

Phase-plane plots of vertical saccades were calculated to depict the trajectory and velocity course of saccades.

*Smooth pursuit and gaze-holding function:* Eye position traces were created after removing noisy saccades or other artifacts (blinks). From this eye position data, eye velocity was calculated by numerical three-point differentiation and subsequent Gaussian low-pass filtering with a corner frequency of 25 Hz. For the Slow Phase Velocity calculation, the high-frequency velocity peaks of nystagmus quick phases, saccades or artifacts were removed from eye velocity with a floating median filter with a time window of 0.5 s. Target and eye velocity traces were then differentiated with a velocity Gaussian low-pass filter of 25 Hz frequency.

Median of smooth pursuit *velocity* gain was calculated as the ratio of eye velocity to target velocity (for simplicity, smooth pursuit gain) measured in the predefined areas. Moreover, *position* smooth pursuit gain, as defined as eye movement amplitude to target amplitude, was calculated to account for eye motility deficits (especially the difference between VSSP and VSGP). Catch-up saccades (fast phases) were removed from gain calculation.

## Statistical analysis

Statistical analysis and figure design were performed using SAS® Software version 9.3 and SPSS® version 22.0.0 (IBM, New York, NY, USA). Graph design was performed by GraphPad® Software version 5.0.3 (La Jolla, CA). Numerical variables in demographic and baseline characteristics were summarized by displaying the following: n (actual sample size), mean, standard deviation, standard error, median, quartiles, 95% confidence intervals, and maximum and minimum. We reported the frequency and percentages (based on the non-missing sample size) of observed levels for all categorical measures. Parametric T-test was used to determine differences in the mean value between NPC patients and HC. Analysis of variance (ANOVA) was used to determine whether a mean value was statistically different among NPC subpopulations (i.e. early-infantile, late-infantile, juvenile, and adults).

To assess which eye movement parameters could predict the severity of patients with NPC (i.e. correlation between VOG impairment and disability scale), we used them as covariates in the univariable linear regression analysis. Moreover, to study whether clinical parameters played a significant role in such correlations, we ran multivariable regression analysis with a stepwise and forward selection method. In linear regression models, R^2^ of the linear regression line including controls are indicated.

All analyses resulted from a single sample of evaluable patients including all of the patients who met the selection criteria and who provided valid information for analysis. Patients who were not physically capable of performing the particular score tasks, or did not perform the test for other reasons, were excluded from the analysis.

## Results

### Clinical state

The disease-specific mDRS score was 10.3 points (SD, 95% CIs of the mean 5.0, [9.4, 11.5]) (n = 69), and SARA was 13.3 points (9.4, [10.9, 15.6]) (n = 63). MoCA was done in 31 patients and resulted in 16.7 points (7.4, [14-19.4]). For the frequency of neurological and psychiatric findings in the study population, see **Table 1**.

### Saccades

For a normal subject as a reference, see **Figure 2A**. As illustrated in **Figures 2B-E**, we observed different patterns of saccadic impairment in patients with NPC. These patterns comprise hypometric saccades (**B**), staircase-pattern (**C**), ‘normal slow’ (**D**), pursuit-like pattern (**E**) (extremely slow, but still movement generated). As a compensating strategy, patients generated blinks to a very significant extent (**F**). The trajectory of saccades can be seen in phase-plane plots of velocity vs. amplitude, see **Figure 3**. For a reference, see normal subject (**3A and 3B**). We observed velocity fluctuation, both in reflexive and self-paced vertical saccades (**3C and 3E**), but not in the horizontal ones (**3D and 3F**).

**Figure 2.**
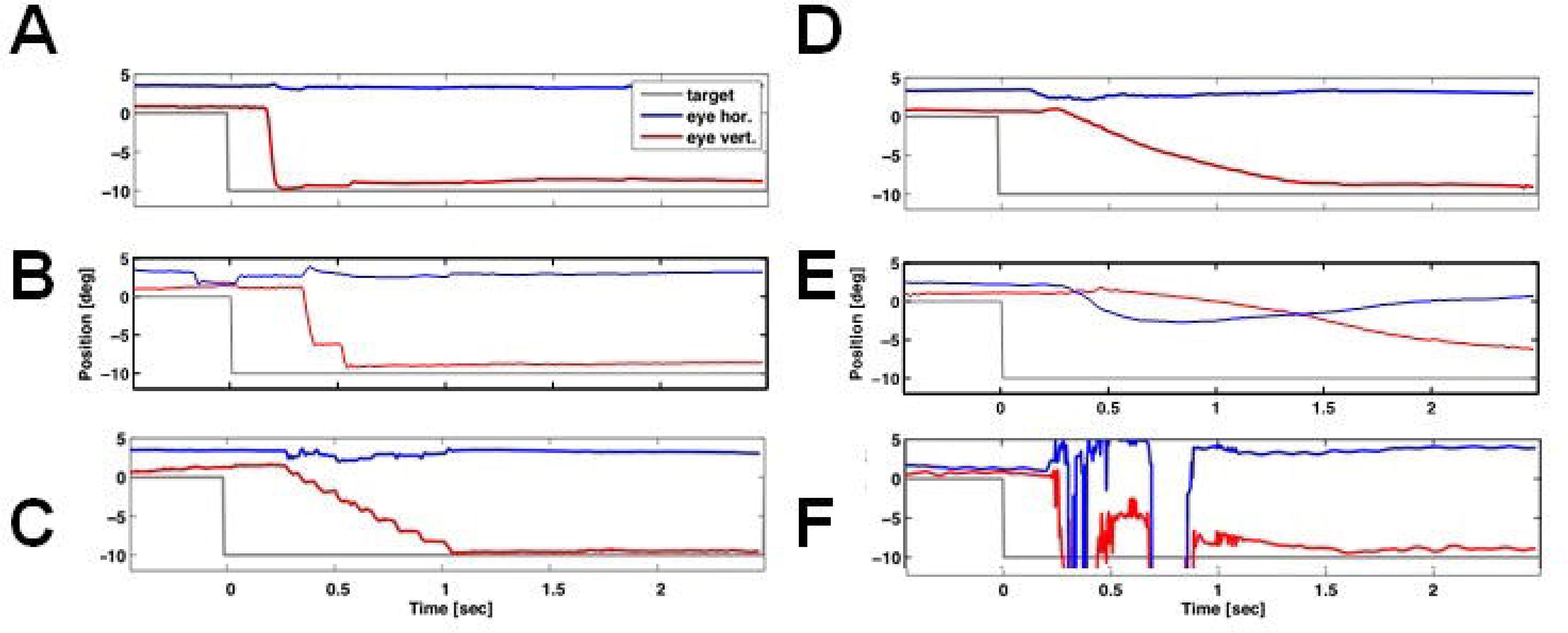
Overview of saccadic phenotypes in a healthy control and in NPC patients: A continuum Fig 2. Phenotypes of vertical downward saccades in NPC disease are depicted. *Red* indicates the vertical eye movement and *blue* indicates the horizontal eye movement. *Grey* depicts the stimulus. In figure A, you see a vertical downward saccade of a normal subject. Note the high peak velocity (steep line) trajectory without fluctuations, latency < 250 ms, amplitude and precision without hypo-or hypermetria, and trajectory without curving. A common finding in NPC is saccadic hypometria (B) that > 20% of the whole saccade. If hypometria is very profound, so-called staircase saccades, seen also often in patients with atypical Parkinson syndromes, are generated (C). Also notice a prolonged latency (time from the stimulus onset to the initiation of the downward movement). Reduced velocity of vertical saccades is depicted in (D), with a *per se* downward saccadic palsy in E. Note that that there is still extremely slow, pursuit-like movement, and the actual saccade performance takes more than two seconds. One of the most remarkable strategies seen in NPC is blinking (F). This strategy is used to initiate saccades that are already impaired due to lack of lid apraxia or blepharospasmus that are typical for parkinsonian disorder. Note the continuum of saccadic impairment with different saccadic phenotypes.

**Figure 3.**
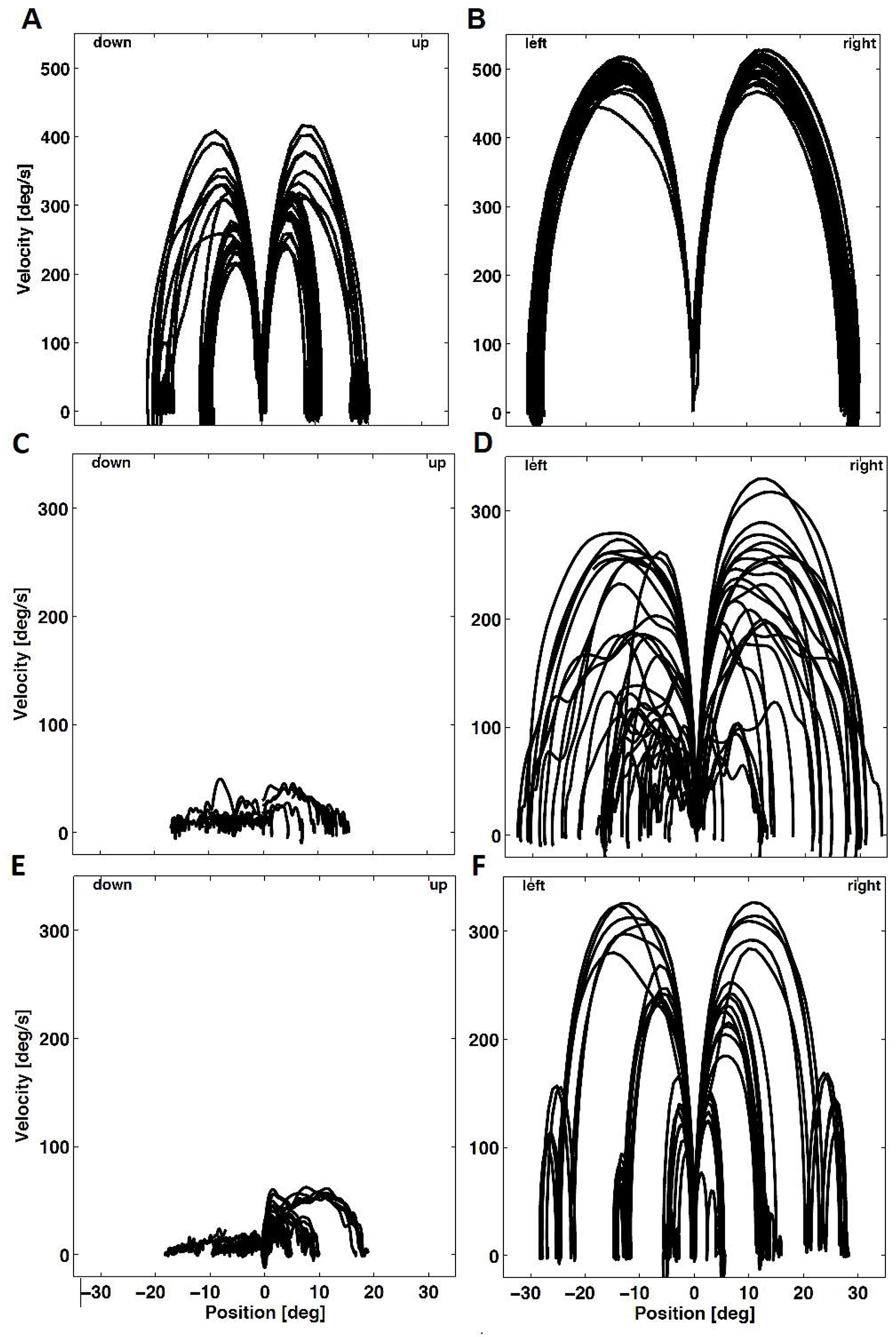
Phase plane plot of vertical saccades in a normal subject and in a patient with Niemann-Pick type C Fig. 3 Phase-plane plots depicting the saccadic amplitude vs velocity. The origin of each saccade has been moved to zero and the velocity at each position along the saccade trajectory is shown. Since larger amplitude saccades typically generate faster velocities, this generates a family of curves where the saccade amplitude can be seen at the endpoint position. Note also the velocity fluctuation of saccades, as shown in the phase-plane plot. Downward saccades tend to be slower, as can be seen in in figure E.

*Reflexive vertical saccades:* Sixty-one patients were able to perform vertical saccades (73.2%) of 10° amplitude **(Fig. 4-1A, C, E, G**), and 58 patients generated saccades to 20° (**Fig. 4-1B, D, F, H)**. Mean PV (for purpose of comprehensiveness, short PV) in response to stimulus of 20° was 63.5°/s (59.5, [47.9-79.2]) in NPC patients and 403.1°/s (69.0, [392.0-414.2°/s]) in healthy subjects (HC) *(p* < 0.001) **(4-1B)**. Regarding the direction, downward saccades yielded 51°/s (68.9, [32.7-69.3]), whilst upward saccades measured 78.8°/s (65.9, [60.8-96.8]), respectively (for both: *p* < 0.001). For saccadic gain (saccadic amplitude/stimulus amplitude), see Section “Vertical Saccades and Vertical Smooth Pursuit” below. The overall characteristics of vertical saccades differed when compared with healthy controls at *p* < 0.001 levels, except for the latency (p < 0.05). The linear regression line slope of vertical saccades to 20° stimulus yielded 21.7 (18.1, [17.0-26.4]) in NPC and 1.6 (0.6, [1.5-1.7]) in HC *(p* < 0.001) **(Figure 4-1G and 4-1H)**.

**Figure 4-1, 4-2 and 4-3.**
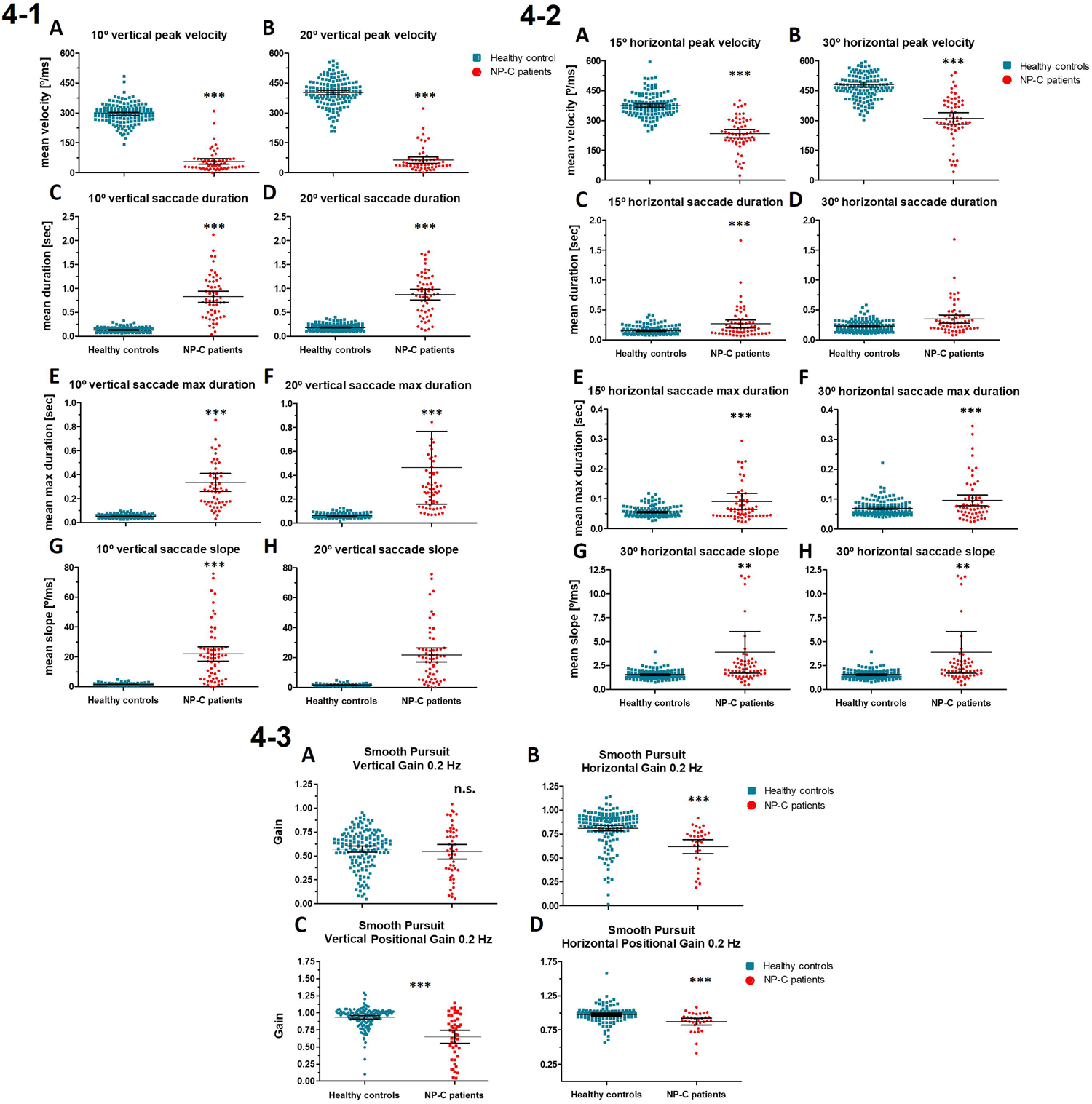
Comparison of vertical and horizontal saccadic parameters and smooth pursuit in patients with Niemann-Pick type C (NPC) and controls Fig. 4 Scatterplot depiction of vertical (4-1), horizontal (4-2) saccadic and smooth pursuit (4-3) parameters. The long vertical line in the scatterplot indicates the mean, the two small horizontal lines indicate the 95% CI. All comparisons, but gain of vertical smooth pursuit 0.2 Hz (4-3A, n.s.), are statistically significant. * p value < 0.05, ** p value < 0.01, *** p value < 0.001

*Reflexive horizontal saccades:* PV of horizontal saccades in response to 30° was 311.7°/s (103.4, [284.6-338.9]) in patients with NPC and 481.7°/s (78.1, [468.8-494.5]) in HC *(p* < 0.001) (for both 15° and 30° saccades, see **Fig. 4-2A-H**). All other saccadic characteristics (amplitude, duration and maximal duration) except latency showed a significant difference. The slope of horizontal saccades was 3.9 (8.4, [1.7-6.1]) in NPC and 1.6 (0.4, [1.5-1.6]) in HC **(Figure 4-2G/H)**. This reflects the lesser impairment of horizontal saccades.

*Self-paced vertical saccades:* PV of upward self-paced saccades to 20° was 158.8°/s (106.2, [39.9, 322]) in NPC patients and 352.7°/s (85.4, [251-459]) in HC (p < 0.001). PV of downward self-paced saccades reached 162.3°/s (146.7, [292.7-106.2]) in NPC, and 352.8°/s (86.4, [458.5-236]) in control group *(p* < 0.001). In contrast to reflexive saccades, NPC patients showed a prolonged intersaccadic interval of self-paced saccades (as an equivalent of latency in reflexive saccades). For upward saccades, this was 2.5 s (1.4, [1.2-2.6]) in NPC, whilst 1.1 s (0.4, [0.8-1.6]) in HC. Regarding the downward saccades, in NPC it was 3.5 s (5.0, [1.2-2.6]) and 1.03 (0.6, [0.8-1.9]) in HC (*p* < 0.001). Patients generated total mean number of 10.6 (19, [2-27]) upward saccades and 8.1 (9.0, [1.0-29.0]) of downward saccades in 30 s, whereas controls generated 30.6 (8.3, [19-41]) upwards and 25.8 (8.7, [16-38]) downwards, respectively (for all *p* < 0.001). Patients achieved a slope at an average of 8.6 (7.8, [1-18]) for upward, and 7.9 (8.1, [1-8.4]) for downward saccades, respectively.

*Self-paced horizontal saccades: PV* of rightward self-paced saccades yielded 244.6°/s (138.6, [179.7-309.5]) in NPC and 449.8°/s (97.2, [432.7-466.8] in HC. To the left, PV reached 240°/s (152.7, [168.5-311.5]) in NPC and 429.8°/s (112.3, [410.1-449.1]) in HC. The intersaccadic interval was, as expected, relevantly prolonged in NPC. To the right, it yielded 2.1 s (1.4, [1.5-2.8]) in NPC and 1.4 s (0.7, [1.2-1.5]) in HC. To the left, the intersaccadic interval was 1.8 s (1.1, [1.3-2.4]) and 1.3 (0.5, [1.2-1.4]) (for both *p* < 0.001). The absolute number of elicited saccades to the right generated in 30 s was 15.7 (8.6, [11.6-19.7]) in NPC and 25.7 (8.7, [24.1-27.2]) in HC (p < 0.001). To the left, the absolute number was 14.6 (8.6, [10.6-18.6]) in NPC and 26.5 (67.3, [91.1-114.8]) in HC, respectively *(p* < 0.001). In all the other saccadic parameters, there were significant differences, when compared to HC *(p* < 0.001, for amplitude *p* < 0.002).

*Reflexive vs. self-paced saccades:* We asked whether there was a significant difference between the two types of examined saccades in NPC. For vertical reflexive saccades, PV was 63.5°/s (59.5, [47.9-79.2]). In contrast, PV of vertical self-paced saccades was 160.6°/s (126.5, [166-214]) (p < 0.001). Interestingly, this was not true for horizontal saccades. For reflexive saccades, mean PV was 311.7°/s (103.4, [284.6-338.9]). In self-paced task, PV reached 242.3°/s (145.7, [174.1-310.5]).

### Smooth pursuit

Vertical smooth pursuit gain in response to 0.2 Hz stimulus frequency was 0.544 (0.269, [0.467-16 0.621] in NPC and 0.572 (0.2, [0.54-0.604]) in HC (p = 0.435) **(Figure 4-3A)**. However, vertical position smooth pursuit gain was 0.649 (0.33, [0.554-0.744]) in NPC and 0.935 (0.149, [0.91-0.959]) in HC *(p* < 0.001) **(Figure 4-3C)**.

Horizontal velocity smooth pursuit gain in response to 0.2 Hz was 0.619 (0.207, [0.546-0.693]) in NPC patients and 0.812 (0.192, [0.781, 0.843]) in HC (p<0.001) **(Figure 4-3B)**. Horizontal position smooth pursuit gain to 0.2 Hz stimulus frequency was 0.979 (0.213, [0.897, 1.060]) in NPC **(Figure 4-3D)**.

### Comparison of vertical saccades and vertical smooth pursuit

98.2% of patients generated vertical saccades (both up and down) that were below the 95% confidence intervals of the controls’ PV. This was not the case for smooth pursuit generated in response to 0.2 Hz stimulation, where only 46.9% of patients had smooth pursuit gain lower than that of 95% of HC. Gain (eye movement amplitude vs stimulus amplitude) of vertical saccades in NPC patients was 0.525 (0.282, [0.451-0.599]) and lower than smooth pursuit gain 0.649 (0.330, [0.554-0.744]) (p = 0.038), while this was not true for HC (vertical saccadic gain: 0.953, (0.112, [0.935-0.971]) vs smooth pursuit gain 0.935, (0.149, [0.910-0.959]; *p* = 0.234) (see also **Fig. 5**).

**Figure 5.**
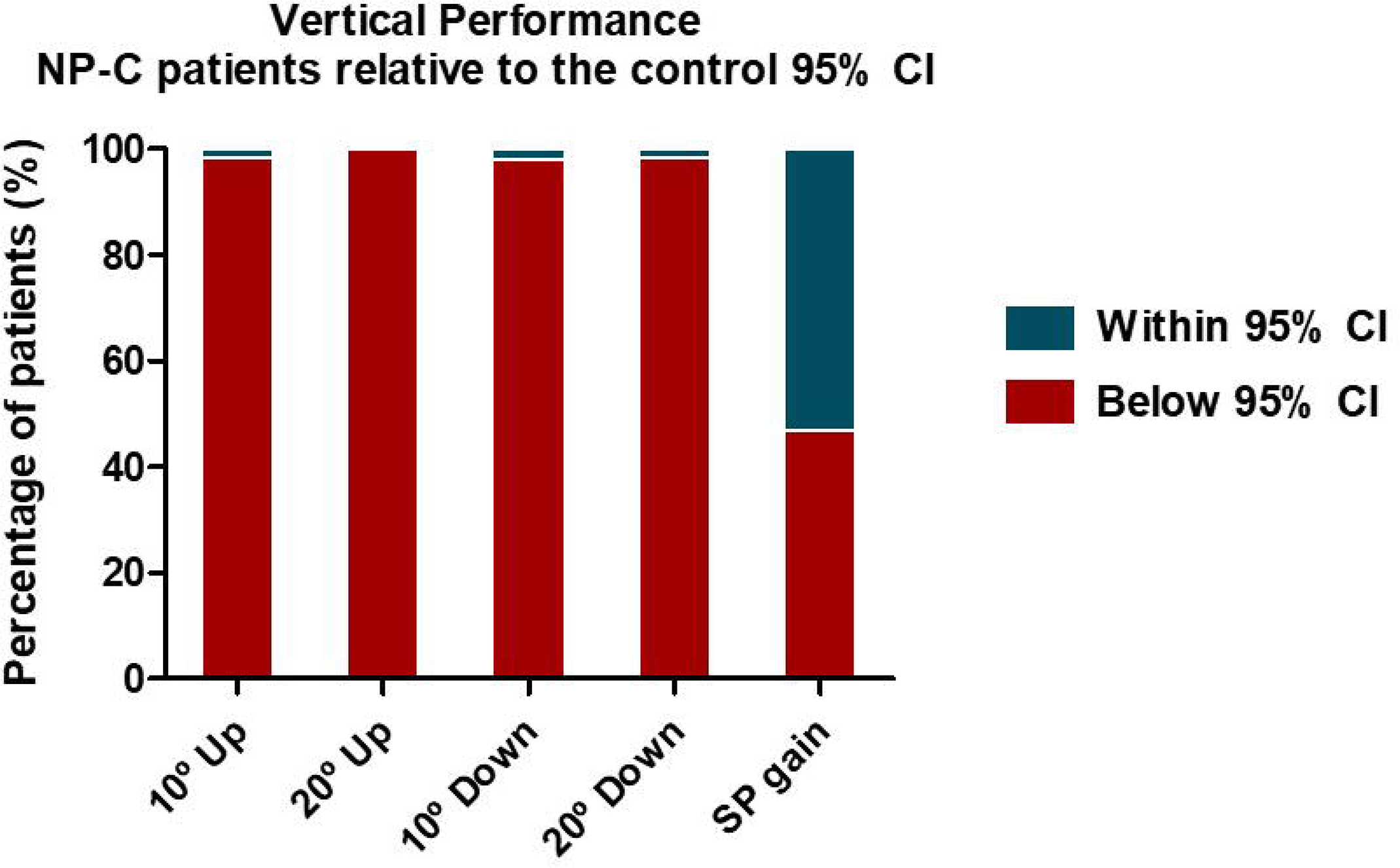
Vertical Supranuclear Gaze vs. Saccade Palsy Fig. 5 Vertical supranuclear saccade palsy (VSSP) was seen in 98.2% of all patients. It was classified as slower than 95% of confidence intervals in the healthy population (red bars). In contrast, only 53.1% of patients demonstrated vertical SGP with slow vertical smooth pursuit. Red bars indicate the values below 95% CI concerning the normal population, green bars indicate the values within 95% CI with regards to normal population.

For a video of a juvenile-onset patient with NPC, suffering VSSP without VSGP, see **Supplemental Video no 1**.

### Gaze-Holding

We did not find a significant gaze-holding deficit in our patient cohort. The number of quick phases, such as saccadic intrusions and quick phases of gaze-evoked nystagmus, was significantly reduced in patients in all five studied positions *(p* < 0.001), when compared to healthy subjects. Vertical peak slow-phase velocity of slow vertical eye drift in all studied positions could be distinguished from that of HC *(p* < 0.001; right *p* < 0.027; down *p* = 0.05). PV of slow horizontal eye drift differed from controls in both eccentric 15° horizontal positions and 10° down, respectively *(p* < 0.001). This was not true for the central position and 10° up.

### Oculomotor characteristics based on age of onset

Due to low number of early-infantile and late-infantile cases is the ANOVA statistical power restricted. The characteristics of 10° saccade did not differentiate based on age of onset. 20° saccades differed in terms of amplitude of both upward and downward saccades, respectively *(p* = 0.035; *p* = 0.022). There was a trend for significance of velocity of upward saccades (*p* = 0.057), and downward amplitude (*p* = 0.051). 20° mean vertical saccadic velocity (*p* = 0.022) and amplitude *(p* = 0.016) were significantly different based on the studied groups. In response to both 15° and 30° stimuli, leftward and rightward saccades did not vary between the studied groups.

Horizontal smooth pursuit and position smooth pursuit varied based on age of onset *(p* < 0.01). This was not true for vertical smooth pursuit.

### Genotype-ocular motor phenotype analysis

Patients with mutations in the sterol-binding domains (n = 12) showed higher PV *(p* = 0.01) and amplitude of vertical saccades *(p* < 0.05), when compared with patients of other genetic backgrounds (*p* < 0.05). For horizontal saccades, there were no statistically relevant differences between ocular motor parameters, based on the mutation-background.

### Univariate linear regression and further analysis

Ocular motor parameters that showed significant relationships with disease-specific and ataxia scales as well as R^2^ values with the tendency to significance are summarized in **Table 2**. All the relationships (VOG parameters vs neurological scales) were plotted and the ones with a significant *p* value and higher R^2^ were evaluated in terms of data dispersion and fit of the regression line.

**Table 2.** Univariant regression analysis of ocular motor parameters and clinical scores. The table shows the exploratory univariant regression analysis of ocular motor parameters and clinical scores. R-squared > 0.5 and in **bold**.

| **SACCADIC PEAK VELOCITY** | **R<sup>2</sup>** |
| --- | --- |
| MDRS Score and 15° horizontal right | 0.41 |
| MDRS Score and 30° horizontal right | 0.37 |
| MDRS Score and 15 ° horizontal left | **0.53** |
| MDRS Score and 30 ° horizontal left | 0.41 |
| MDRS Score and 15 degree horizontal combined left-right | 0.46 |
| MDRS Score and 30 degree horizontal combined left-right | 0.38 |
| SARA Score and 15 degree horizontal right | 0.33 |
| SARA Score and 30 degree horizontal right | 0.31 |
| SARA Score and 15 degree horizontal left | 0.37 |
| SARA Score and 30 degree horizontal left | 0.32 |
| SARA Score and 15 degree horizontal combined left-right | 0.34 |
| SARA Score and 30 degree horizontal combined left-right | 0.31 |
| SCAFI9HPTD Score and 15 degree horizontal right | 0.31 |
| SCAFI9HPTD Score and 15 degree horizontal left | 0.42 |
| SCAFI9HPTD Score and 30 degree horizontal left | 0.35 |
| SCAFI9HPTD Score and 15 degree horizontal combined left-right | 0.35 |
| SCAFI9HPTD Score and 30 degree horizontal combined left-right | 0.28 |
| SCAFI9HPTN Score and 15 degree horizontal left | 0.37 |
| SCAFI9HPTN Score and 30 degree horizontal left | 0.35 |
| SCAFI9HPTN Score and 15 degree horizontal combined left-right | 0.3 |
| SCAFI9HPTN Score and 30 degree horizontal combined left-right | 0.28 |
| SCAFIPATA Score and 15 degree horizontal left | 0.29 |
| SCAFIPATA Score and 30 degree horizontal left | 0.28 |
| **LATENCY** |  |
| SCAFI9HPTD Score and 15 degree horizontal right | **0.56** |
| SCAFI9HPTD Score and 30 degree horizontal right | 0.34 |
| SCAFI9HPTD Score and 30 degree horizontal left | 0.49 |
| SCAFI9HPTD Score and 15 degree horizontal combined left-right | 0.32 |
| SCAFI9HPTD Score and 30 degree horizontal combined left-right | 0.43 |
| SCAFI9HPTN Score and 15 degree horizontal right | 0.37 |
| SCAFI9HPTN Score and 30 degree horizontal left | 0.33 |
| SCAFI9HPTN Score and 15 degree horizontal combined left-right | 0.29 |
| SCAFI9HPTD Score and 10 degree vertical up | 0.4 |
| SCAFI9HPTD Score and 10 degree vertical down | 0.41 |
| SCAFI9HPTD Score and 10 degree vertical combined up-down | 0.37 |
| SCAFI9HPTN Score and 10 degree vertical up | 0.35 |
| SCAFI9HPTN Score and 10 degree vertical down | 0.38 |
| SCAFI9HPTN Score and 10 degree vertical combined up-down | 0.33 |
| AMPLITUDE |  |
| MDRS Score and 10 degree vertical up | 0.34 |
| **MDRS Score and 20 degree vertical up** | **0.56** |
| MDRS Score and 10 degree vertical down | 0.41 |
| **MDRS Score and 20 degree vertical down** | **0.61** |
| MDRS Score and 10 degree vertical combined up-down | 0.35 |
| **MDRS Score and 20 degree vertical combined up-down** | **0.58** |
| SARA Score and 20 degree vertical up | 0.34 |
| SARA Score and 20 degree vertical down | 0.38 |
| SARA Score and 20 degree vertical combined up-down | 0.36 |
| SCAFI9HPTD Score and 10 degree vertical combined up-down | 0.32 |
| SCAFI9HPTD Score and 20 degree vertical combined up-down | 0.34 |
| SCAFIPATA Score and 20 degree vertical up | 0.49 |
| SCAFIPATA Score and 20 degree vertical down | 0.31 |
| SCAFIPATA Score and 20 degree vertical combined up-down | 0.38 |
| MDRS Score and 20 degree vertical up | 0.21 |
| MDRS Score and 20 degree vertical down | 0.21 |
| MDRS Score and 20 degree vertical combined up-down | 0.21 |
| DURATION |  |
| MDRS Score and 10 degree vertical up | 0.48 |
| MDRS Score and 20 degree vertical up | 0.41 |
| MDRS Score and 10 degree vertical down | **0.68** |
| MDRS Score and 20 degree vertical down | **0.55** |
| MDRS Score and 10 degree vertical combined up-down | **0.57** |
| MDRS Score and 20 degree vertical combined up-down | 0.46 |
| SARA Score and 10 degree vertical up | 0.44 |
| SARA Score and 20 degree vertical up | 0.35 |
| SARA Score and 10 degree vertical down | **0.56** |
| SARA Score and 20 degree vertical down | 0.47 |
| SARA Score and 10 degree vertical combined up-down | 0.49 |
| SARA Score and 20 degree vertical combined up-down | 0.41 |
| MDRS Score and Position Gain vertical Freq 0.2 Hz | 0.32 |
| SARA Score and Position Gain vertical Freq 0.2 Hz | 0.31 |

Univariate linear regression analysis showed that duration of 10° saccades is related to disease state (R^2^ = 0.68) (**Fig. 6 A and 6B**). This especially concerning 10° vertical down (**Table 2**). 20° vertical down amplitude tracked with mDRS (R^2^ = 0.61) (**Fig. 6 C)**. 20° vertical saccadic amplitude was a negative predictor to time of SCAFI 9HPTD (R^2^ = 0.58) (**Fig. 6 D**), but also mDRS (R^2^ = 0.58). PV of vertical saccades was not useful in predicting disease severity (for up/down and 10°/20°, R^2^ <0.2).

**Figure 6.**
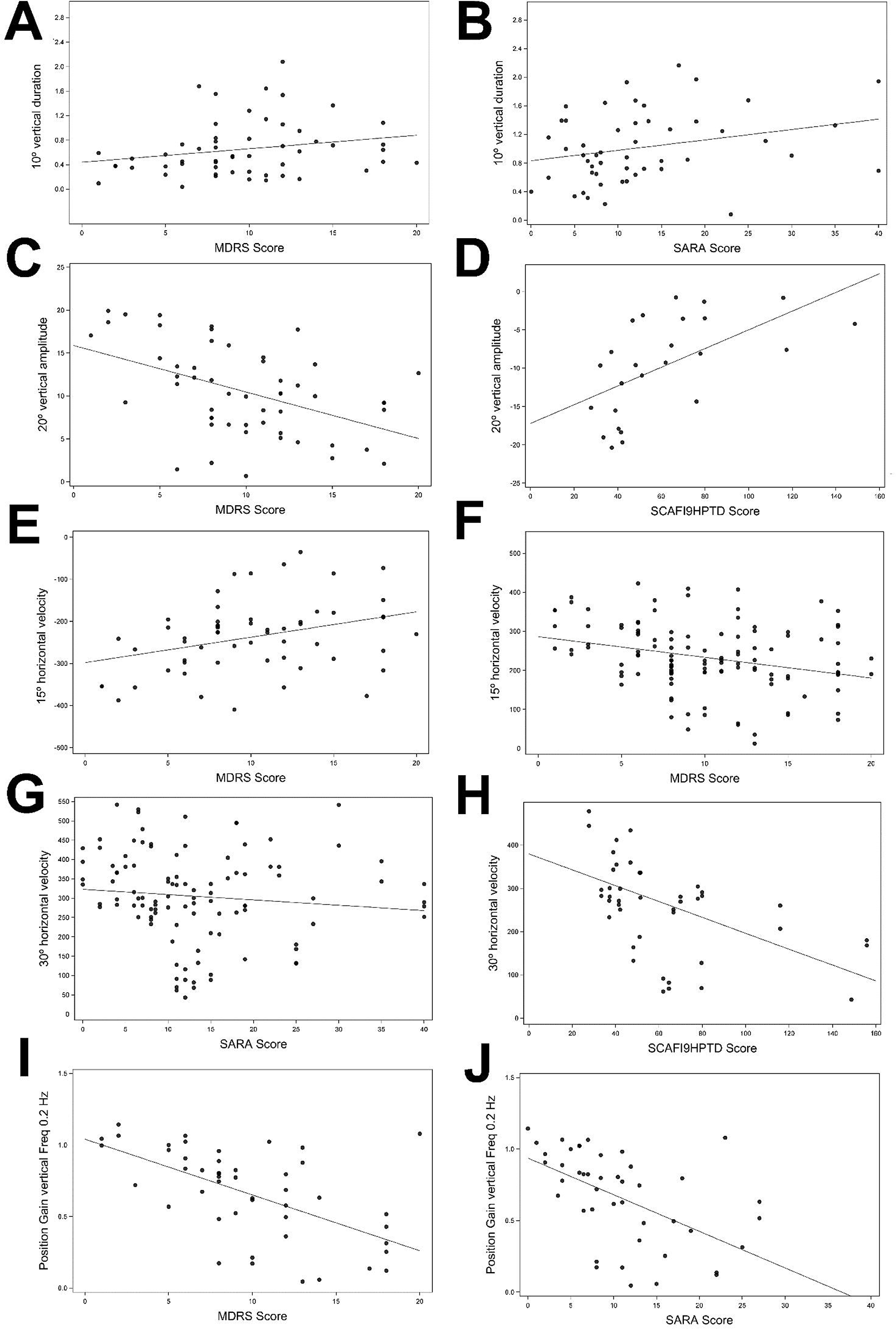
Saccadic and smooth pursuit parameters and their relationships to neurological scores

PV of 15° left horizontal saccades was significantly related to mDRS scale (R^2^ = 0.53) (**Fig. 5 E)**, when combined 15° horizontal left-right PV (R^2^ = 0.46) (**Fig. 6 F)**. Horizontal saccadic combined (left-right) PV and SARA score yielded R^2^ of 0.38 (30° saccades) and R^2^ of 0.37 (15° saccades), respectively **(Fig. 6 G)**. SCAFI domain 9HPTD and PV of 15° left-right saccades yielded R^2^ of 0.35 (**Fig. 6 H**). Latency of 15° rightward saccades tracked with SCAFI 9HPTD Score (R^2^ = 0.56).

Position smooth pursuit gain to 0.2 Hz stimulation was related to both mDRS and SARA scores to a modest extent (R^2^ = 0.32 vs R^2^ = 0.31) (**Fig. 6I** and **6J**). Gaze-holding did not reflect the disease-severity.

Miglustat treatment was used as a covariate in the multivariate regression analysis; however, the assessment of its role was hampered by the patient-to-patient variability and restricted sample sizes.

PV of 10° upward saccades (p = 0.046) in treated >1 year was significantly higher (with a significantly higher slope), probably due to NPC disease duration, *de facto* suffering an advanced disease. Regarding horizontal saccades, patients treated for >1y exhibited the following characteristics: lower peak velocity to 15° stimulus (right *p* = 0.048; left *p* = 0.031) and to 30° stimulus *(p* = 0.004), respectively. Further, higher 30° slope *(p* = 0.008), 15° left amplitude *(p* = 0.044), 30° left peak velocity (p = 0.05) and 30° left amplitude *(p* = 0.008) were noted.

When considering other variables, disease duration played a significant role concerning ocular motor function.

## Discussion

Despite the fact that the hallmark of the NPC disease is ocular motor impairment, ocular motor function in NPC disease has not been systematically analyzed in a large cohort of patients due to its rarity and underdiagnosis. Thus, in this study, we systematically examined a range of ocular motor functions, namely reflexive and self-paced vertical and horizontal saccades, smooth pursuit and gaze-holding as functional outputs of the brainstem, cerebellum and cortical centers. The major findings of this multi-center cross-sectional prospective study are as follows:

*First*, quantitative measurements of eye movements showed that vertical supranuclear saccade function, not vertical gaze palsy, is the cardinal ocular motor sign of NPC disease. *Second*, contrary to what is commonly reported, downward *and* upward saccades are impaired to a similar extent. *Third*, reflexive vertical saccades were more impaired and slower than self-paced ones. In contrast, the velocity of horizontal self-paced saccades was lower than that of their reflexive counterparts. *Fourth*, horizontal saccadic peak velocity and latency, as well as amplitude and duration of vertical saccades were the most relevant variables to consider reflecting the clinical state. Saccades of smaller amplitudes are more useful than the larger ones in both vertical and horizontal planes. This suggests that they can be utilized as useful surrogate indicators for severity. *Fifth*, patients apply blinking and head and upper body movements to initiate and accelerate the saccades. Clinicians should actively search for these clues. *Sixth*, the ocular motor status was dependent on certain covariates, such as age of onset and disease duration. *Seventh*, vertical saccades of patients with juvenile forms were more constrained than that of adult patients. *Eight*, we observed patient specific-saccadic phenotypes, but due to the compound heterozygosity, the genotype-ocular motor phenotype correlation has not been possible. *Ninth*, the ocular motor profiling with regards to genetic background showed that in 12 patients with mutations in the sterol-sensing domain (transmembrane helices 3-8, amino acides 616-854) there was a higher peak velocity of vertical saccades and larger saccadic vertical amplitude, when compared with other groups.

Finally, we observed no significant effect of treatment with miglustat in terms of ocular motor function, likely due to insufficient information about treatment duration; nevertheless, we hypothesize that there was a therapy bias due to severely disabled patients, diagnosed and suffering the disease longer and thus, having been treated longer with miglustat.

### Vertical Supranuclear Saccadic vs. Gaze Palsy

Up to now, the VSGP, but not VSSP has been highlighted in the literature as a cardinal symptom of NPC, not acknowledging the actual ocular motor impairment. Since in clinical practice the ocular motor examination is often restricted to following the target, when examining smooth pursuit, the slow vertical saccades can be overlooked. Thus, through a thorough ocular motor examination, focusing on both saccades and smooth pursuit, clinicians can detect VSSP. VSGP has been shown to be present in 70% of patients from the international patient registry (Patterson *et al*., 2013). However, here we show that VSSP is present in 98.2% of all patients, but VSGP in only 46.9%. This corresponds with the comparison of saccadic and smooth pursuit gains, which showed that motility is more restricted in vertical saccades than in smooth pursuit. VSGP is present in the advanced stages of ocular motor impairment, which is at a point where, based on the symptomatology, the disease is often already recognized. Nevertheless, for early diagnosis and treatment, VSSP is crucial in this neurodegenerative disease, as otherwise the window of opportunity for a possible treatment response is lost.

Vertical saccades are often accompanied by “around the house” sign (curved saccade trajectory when plotted in X-Y space), which is typical of saccadic palsy in one respective plane (Eggink *et al*., 2016), and is also seen in other lysosomal storage diseases, such as Gaucher disease type 3 (Bremova-Ertl *et al*., 2017). Our study reiterates that the hallmark of NPC disease is VSSP and not VSGP, as you can see in the Supplemental Video no S1. Thus, clinicians should be trained to recognize and differentiate impairments of the saccadic system from impairments of smooth pursuit, and to understand the possibility of combined deficits. To emphasize the importance of the difference between VSSP and VSGP, we suggest revising of the current terminology with regards to saccadic vs. gaze palsy also in other inborn errors of metabolism, such as Gaucher disease type 3 (GD3) and neurodegenerative diseases, such as progressive supranuclear palsy (PSP). Saccadic palsy is the initial symptom due to the selective vulnerability of the saccadic neurons, probably due to their high metabolic properties, but the underlying pathophysiology is not known.

### Vertical saccadic patterns and VSSP/VSGP compensating strategies

We observed different saccadic patterns that tended to be patient-specific. We recognized slow saccades, and extremely slow, “pursuit-like” saccades, where the saccade resembled a pursuit movement. The question was raised whether the movement was still generated by burst neurons in riMLF, or whether the pursuit system also played a role. Saccade-generating burst neurons are inhibited by omnipause neurons, except during saccadic performance and blinks (Rucker *et al*., 2004, 2011). Due to transient velocity fluctuations during the saccade, suggestive of omnipause neuron dysfunction, we argue for the former. Further, patients generated hypometric saccades with either one large corrective saccade (>20% of amplitude of first saccade), or so-called stair-case pattern saccades. The number of saccadic steps varied from two to 10. These highly hypometric saccades are common for atypical Parkinson syndromes, such as PSP, the main differential Bremova-Ertl et. al, 2020 diagnosis for the adult form of NPC. This further establishes the link between NPC and other neurodegenerative disorders (Bhidayasiri *et al*., 2001).

We emphasize that compensating strategies patients use in the daily life to overcome the ocular motor deficits can also add to establishing the diagnosis. To elicit the saccades, patients tend to blink in order to silent omnipause neurons that inhibit burst excitatory neurons (BEN). To overcome the palsy of gaze, patients produce high-amplitude head movements, or even upper body movements. This can be often seen also in children (Bremova and Strupp, 2017). We strongly encourage the clinicians to actively look for these movement clues that compensate for subclinical ocular motor impairment and should prompt a thorough and detailed ocular motor, but neurological examination, respectively. It is important to note that in order to produce valid video-oculographic data, saccades that are confounded by blinks and/or linked to pathological head/upper body movements, must be excluded, as in our study.

### Reflexive vs. Self-paced saccades

Peak velocity of vertical self-paced and reflexive saccades differed to a relevant extent whereas the horizontal saccades did not show a significant difference. This is surprising, since BEN for both modalities have a common supranuclear center, namely riMLF for vertical and pontine paramedian reticular formation (PPRF) for the horizontal saccades, respectively (Buttner-Ennever, 2008). The firing rate of the BEN is linearly aligned with the saccadic velocity (Van Gisbergen *et al*., 1981). The reason why self-paced saccades reach higher peak velocity is not clear, especially reflecting cognition and processing speed also in other degenerative and traumatic brain conditions (Ettenhofer and Barry, 2016; Taghdiri *et al*., 2018; Hunfalvay *et al*., 2019).

There might be a difference in involvement of supra-riMLF pathways, namely frontal eye field (FEF) for self-paced and parietal eye field (PEF) for reflexive saccades. The characteristics of both self-paced and reflexive saccades were pathological, when compared to healthy population. We speculate that the rapidly changing visual stimulus for the highly disturbed vertical reflexive saccades might represent a relevant stressor that compromise the performance. Horizontal saccades showed also a remarkable decrease of peak velocity and prolongation of duration, showing the functional impairment of PPRF in pons, but since they are affected to a lesser extent, they are easier for patients to perform regardless the visual task.

As expected, latency in vertical reflexive saccades was normal; however, the intersaccadic interval was greatly prolonged. This reiterates the progressive frontal impairment in patients with NPC. Considering disease progression and later involvement of smooth pursuit, with vertical being more affected than horizontal, ocular motor brainstem and cerebral regions are affected earlier in the course of disease, followed by the cerebellar ocular motor abnormalities (even though saccadic Bremova-Ertl et. al, 2020 gain is also highly dependent on the cerebellum). This corresponds to the findings of a prior neuropathological study (Solomon *et al*., 2005).

### Upward vs. Downward saccades: No Significant Differences Noted

In contrast to the common believed discrepancy between the downward and upward saccades due to the mono-(ipsi-) and bilateral supranuclear innervation of oculomotor nuclei by the riMLF (Horn and Buttner-Ennever, 1998), both down- and upward saccades showed similar characteristics, including peak velocities. This might be caused by the fact that the bilateral upward innervation is counteracted by the gravity. In neurodegeneration, the same degree of functional impairment results in both directions. Also, the less frequent upward eye movements might lead to a worse upward motor function, when compared with common downward eye movements, as the result of a training-effect.

### Potential biomarkers to use in clinical trials, further significance and result exploitation

We asked which ocular motor parameters track with the NPC disease to the greatest extent. The characteristics of smaller, 15° horizontal saccades, especially amplitude, but also velocity, duration and latency reflected well the state of disease, quantified by clinical scores. This is also the case in vertical saccades, where smaller, 10° amplitude saccades also more sensitive in terms of disease severity. The reason, why 20° vertical saccades are not useful is the very often blinking to initiate and speed-up these high-acceleration, large-amplitude saccades, thus confounding them. On the other hand, the fact that the amplitudes of these large saccades often tend to be hypometric, can make at least some use of them, once the eye lids are strained.

Even though vertical saccades in both directions show similar peak velocities, the duration of downward saccades is a more sensitive parameter in terms of the state of disease.

Our results suggest, that position, not velocity smooth pursuit profile can be established as surrogate parameters. Interestingly, this is more the case for vertical, not horizontal smooth pursuit.

The characteristics of self-paced saccades might be a useful biomarker, especially total count and peak velocity. Further ocular motor studies using this task are needed. In terms of important covariants, disease duration affected the ocular motor systems to a considerable extent, as was expected.

This once more establishes the fact that vertical saccades deteriorate to the level that makes them unusable as clinical biomarkers, especially because they are often not eligible for clinical trials due to severe deterioration at the time of diagnosis.

Bremova-Ertl et. al, 2020 Considering the further importance of our findings, ocular motor function in NPC1 heterozygotes (one mutation carriers) is similar to patients with manifest disease (with two underlying mutations), replicating the neurodegenerative pattern (Bremova-Ertl *et al*., 2020). Thus, the identified ocular motor parameters can be also used in NPC1 heterozygotes to screen for neurological impairments, especially with regards to Parkinson phenotype. In addition, once treated, also to evaluate the disease progression and the effects of treatment.

### Age of Onset

With regards to the type of NPC disease based on age of onset, vertical saccades of patients with juvenile forms showed the most pronounced deficits, the comparison of infantile patients’ data were restricted of the small sample size. This was also true for other ocular motor systems, including horizontal pursuit movements. This suggests that there is a specific pattern of neurodegeneration in the brainstem and the cerebellum based on the time of disease onset.

### Gaze-evoked nystagmus

We did not find a clear gaze-holding nystagmus. This is not surprising in the light of the fact that the saccadic system is most impaired in this disease. The number of quick phases was also reduced when compared with normal population. This reinforces the fact that patients suffer predominant saccadic deficits. We observed an ocular motor drift vertically that was simply not followed by a saccade. Since cerebellar ataxia, a clinical expression of common cerebellar atrophy (Solomon *et al*, 2005; Abel *et al*, 2012; Fusco *et al*, 2012; Bowman *et al*., 2015), is often present, the fixation instability even without observation of gaze-holding nystagmus is suggestive of functional impairment of vestibulo-cerebellum in later stages of disease, and also of neural integrator InC (Crawford *et al*., 1991).

### The role of miglustat

The only currently available disease-stabilizing therapy is the treatment with miglustat (Zavesca®) (Patterson *et al*., 2007). Other treatment options were recently evaluated in clinical trials, including Hydroxypropyl Beta Cyclodextrin (VTS-270) NCT01747135 and Arimoclomol NCT02612129. The results of the former trial were negative (Sidhu *et al*., 2020), whilst the latter was able to show a significant therapeutic benefit (Kirkegaard *et al*., 2016). A multicenter, international open label, phase II study to evaluate the efficacy and safety of N-acetyl-L-leucine in patients (IB1000) with NPC (NCT03759639) is underway.

The limitations of the currently available treatment modalities stresses the importance of establishing the diagnosis in the early stages of the disease. This may reduce or stagnate neurological progression and prolong life-expectancy among patients. In addition, the disease may be modified by employing one or more of the available CNS-active medications, which influence the ocular motor function both positively and negatively (Noachtar *et al*., 1998, Schillevoort *et al*., 2001*b*, *a*).

Regarding the retrospective evaluation of the treatment with miglustat, we did not find a profound therapeutic benefit regarding ocular motor function. Interestingly, when we divided patients into two groups on medication and without the medication, we found that patients treated for more than one year featured more profound saccadic deficits. When the miglustat treatment was taken as a covariate in the model, it turned out to be irrelevant in terms of ocular motor function, likely because patients treated for more than one year also suffered from a more advanced stage of disease. Nevertheless, this finding cannot be generalized due to the cross-sectional, non-controlled study design.

### Genotype-ocular motor phenotype relationship

To our knowledge, this is a first attempt to address the question of whether certain mutations affect the ocular motor system more profoundly than others. Regarding the disease state, there is evidence that some specific mutations are more severe, leading to greater lipid storage (Fusco *et al*., 2012; Xiong *et al*., 2012). This data suggests a differential effect of mutation group and that more severe biochemical deficits may correlate to higher degrees of ocular motor impairment. However, our findings are limited by sample size and must be interpreted with caution due to compound heterozygosity. In addition, some mutations lead to no protein production, and this effect cannot be allocated to a protein domain (but these patients are usually severe affected, thus being not able to perform the video-oculographic task).

### Limitations

The primary limitation of this study is clearly the disease complexity *per se* and the level of the patients’ disability, which leads to missing data. Further, dividing the patient population into groups leads to smaller sample sizes, thus reducing the power of the study. Also, the cross-sectional and uncontrolled design of the study is a limitation in this progressive neurodegenerative disease. Our next step is to assess the ocular motor systems of a large cohort of NPC patients over a period longer than six months. This might shed light on the disease progression and on the influence of lipid accumulation on ocular motor function and specific ocular motor phenotypes.

## Conclusions

In contrast to what is commonly believed and reported in the literature, the cardinal ocular motor sign of the NPC disease is the vertical supranuclear *saccade* palsy present in 98.2% of 72 patients we examined, whereas gaze palsy was found in only 46.9%. We emphasize that all patients with VSGP demonstrate a saccadic impairment, but not all patients with VSSP feature the gaze palsy. Both downward and upward saccades are affected to the same extent.

The parameter that could serve as the surrogate endpoint is peak velocity of horizontal saccades, but other horizontal saccade characteristics can also be used. Further, position, but not velocity-based vertical smooth pursuit gain and vertical saccadic duration can be taken as the further secondary endpoints. We recommend examining horizontal saccades and vertical smooth pursuit to track disease progression and to evaluate the efficacy of administered therapies both in clinical trials and in clinical practice.

VSSP can help clinicians diagnose the disease at the initial stages. It is important to diagnose the disease in the early phases to initiate disease-modifying therapy. This may be able to slow down neurologic progression and thus, prolong the life-expectancy of affected individuals.

All in all, this study is relevant not just with regards to NPC disease, but also a number of other neurodegenerative and IEM diseases with neuro-ophthalmological manifestations.

## Acknowledgments

We thank R. John Leigh for his critical review and input. We thank our collaborators Matthis Synofzik, MD, PhD and Hans Kluenemann MD^12^ for helping us with the study. We thank Sarah Hoffmann, MD for helping us with the study, Katie Göttlinger and Adriana Brueggemann, MD for copyediting the manuscript. This study was supported by Actelion, Ltd, a Janssen company of Johnson & Johnsons.

## Author contribution

Dr. T. Bremova-Ertl: design, data collection, data interpretation and discussion of the data, writing of the manuscript. Dr. L. Abel and Dr. Walterfang: data collection and revising the manuscript for the important intellectual content. Dr. E. Salsano: discussion of the data, patient recruiting and revising the manuscript for the important intellectual content. Dr. A. Ardissone: patient recruiting and revising of the manuscript. Dr. V. Malinova: patient recruitment, revising of the manuscript. Dr. M. Kolnikova: patient recruiting and revising of the manuscript. Dr. J. Gascon Bayarri: data collection and revising the manuscript. Dr. A. Tavasoli and Dr. M. Ashrafi: data collection and revising the manuscript for the important intellectual content. Dr. Y. Amraoui: patient recruiting and revising of the manuscript. Dr. E. Mengel: patient recruiting and revising of the manuscript for the important intellectual content. Dr. S. Kolb and Dr. A. Brecht: general support and revising of the manuscript. Dipl.-Ing. S. Bardins: design, data analysis. Dr. M. Strupp: idea of the study, discussion of the data, revising of the manuscript.

## References

Abel LA, Bowman EA, Velakoulis D, Fahey MC, Desmond P, Macfarlane MD, et al. Saccadic eye movement characteristics in adult Niemann-Pick Type C disease: relationships with disease severity and brain structural measures. PloS One 2012; 7: e50947.

Abel LA, Walterfang M, Fietz M, Bowman EA, Velakoulis D. Saccades in adult Niemann-Pick disease type C reflect frontal, brainstem, and biochemical deficits. Neurology 2009; 72: 1083–1086.

Abel LA, Walterfang M, Stainer MJ, Bowman EA, Velakoulis D. Longitudinal assessment of reflexive and volitional saccades in Niemann-Pick Type C disease during treatment with miglustat. Orphanet J Rare Dis 2015; 10: 160.

Bahill A, Clark MR, Stark L. The main sequence, a tool for studying human eye movements. Math Biosc 1975;24:191–204.

Bhidayasiri R, Riley DE, Somers JT, Lerner AJ, Büttner-Ennever JA, Leigh RJ. Pathophysiology of slow vertical saccades in progressive supranuclear palsy. Neurology 2001; 57: 2070–2077.

Bowman EA, Walterfang M, Abel L, Desmond P, Fahey M, Velakoulis D. Longitudinal changes in cerebellar and subcortical volumes in adult-onset Niemann-Pick disease type C patients treated with miglustat. J. Neurol. 2015; 262: 2106–2114.

Bremova T, Krafczyk S, Bardins S, Mengel E, Reinke J, Strupp M. Vestibular function in Niemann-Pick type C. J Neurol. 2016; 263: 2260–2270.

Bremova T, Malinová V, Amraoui Y, Mengel E, Reinke J, Kolníková M, et al. Acetyl-dl-leucine in Niemann-Pick type C: A case series. Neurology 2015; 85: 1368–1375.

Bremova T, Strupp M. Vertical Supranuclear Gaze Palsy in a Toddler With Niemann-Pick Type C. Pediatr Neurol. 2017; 72:94.

Bremova-Ertl T, Schiffmann R, Patterson MC, Belmatoug N, Billette de Villemeur T, Bardins S, et al. Oculomotor and Vestibular Findings in Gaucher Disease Type 3 and Their Correlation with Neurological Findings. Front Neurol 2018; 15: 8:711.

Bremova-Ertl T, Sztatecsny C, Brendel M, Moser M, Möller B, Clevert DA, et al. Clinical, ocular motor, and imaging profile of Niemann-Pick type C heterozygosity. Neurology 2020 21; 94: e1702–e1715.

Buttner-Ennever JA. Mapping the oculomotor system. Prog Brain Res 2008; 171: 3–11.

Büttner-Ennever JA, Eberhorn A, Horn AKE. Motor and sensory innervation of extraocular eye muscles. Ann N Y Acad Sci 2003; 1004: 40–49.

Büttner-Ennever JA, Horn AKE. The neuroanatomical basis of oculomotor disorders: the dual motor control of extraocular muscles and its possible role in proprioception. Curr Opin Neurol 2002; 15: 35–43.

Chen AL, Riley DE, King SA, Joshi AC, Serra A, Liao K, et al. The Disturbance of Gaze in Progressive Supranuclear Palsy: Implications for Pathogenesis. Front Neurol 2010 3; 1:147.

Crawford JD, Cadera W, Vilis T. Generation of torsional and vertical eye position signals by the interstitial nucleus of Cajal. Science 1991; 252: 1551–1553.

Eggink H, Brandsma R, van der Hoeven JH, Lange F, de Koning TJ, Tijssen MAJ. Teaching Video NeuroImages: The ‘round the houses’ sign as a clinical clue for Niemann-Pick disease type C. Neurology 2016 May 10; 86: e202.

Ettenhofer ML, Barry DM. Saccadic Impairment Associated With Remote History of Mild Traumatic Brain Injury. J Neuropsychiatry Clin Neurosci 2016; 28: 223–31.

Fusco C, Russo A, Galla D, Hladnik U, Frattini D, Giustina ED. New Niemann-Pick Type C1 Gene Mutation Associated With Very Severe Disease Course and Marked Early Cerebellar Vermis Atrophy. J Child Neurol 2013; 28: 1694–7.

Garbutt S, Harwood MR, Kumar AN, Han YH, Leigh RJ. Evaluating small eye movements in patients with saccadic palsies. Ann N Y Acad Sci 2003; 1004: 337–346.

Garver WS, Francis GA, Jelinek D, Shepherd G, Flynn J, Castro G, et al. The National Niemann-Pick C1 disease database: report of clinical features and health problems. Am. J. Med. Genet. A 2007; 143A: 1204–1211.

Havla J, Moser M, Sztatecsny C, Lotz-Havla AS, Maier EM, Hizli B, et al. Retinal axonal degeneration in Niemann-Pick type C disease. J Neurol. 2020 Mar 28.

Horn AK, Buttner-Ennever JA. Premotor neurons for vertical eye movements in the rostral mesencephalon of monkey and human: histologic identification by parvalbumin immunostaining. JComp Neurol 1998; 392: 413–427.

Hunfalvay M, Roberts C-M, Murray N, Tyagi A, Kelly H, Bolte T. Horizontal and vertical self-paced saccades as a diagnostic marker of traumatic brain injury. Concussion Lond Engl 2019; 4: CNC60.

Iturriaga C, Pineda M, Fernández-Valero EM, Vanier MT, Coll MJ. Niemann-Pick C disease in Spain: clinical spectrum and development of a disability scale. J Neurol Sci 2006; 249: 1–6.

Kirkegaard T, Gray J, Priestman DA, Wallom K-L, Atkins J, Olsen OD, et al. Heat shock protein-based therapy as a potential candidate for treating the sphingolipidoses. Sci Transl Med 2016; 8: 355–118.

Leigh RJ, Kennard C. Using saccades as a research tool in the clinical neurosciences. Brain 2004; 127: 460–477.

Nasreddine ZS, Phillips NA, Bédirian V, Charbonneau S, Whitehead V, Collin I, et al. The Montreal Cognitive Assessment, MoCA: a brief screening tool for mild cognitive impairment. J Am Geriatr Soc 2005; 53: 695–699.

Noachtar S, von Maydell B, Fuhry L, Buttner U. Gabapentin and carbamazepine affect eye movements and posture control differently: a placebo-controlled investigation of acute CNS side effects in healthy volunteers. Epilepsy Res 1998; 31: 47–57.

Patterson MC, Mengel E, Vanier MT, Schwierin B, Muller A, et al. Stable or improved neurological manifestations during miglustat therapy in patients from the international disease registry for Niemann-Pick disease type C: an observational cohort study. Orphanet Journal of Rare Diseases 2015

Patterson MC, Mengel E, Wijburg FA, Muller A, Schwierin B, Drevon H, et al. Disease and patient characteristics in NPC patients: findings from an international disease registry. Orphanet J Rare Dis 2013; 8: 12.

Patterson MC, Clayton P, Gissen P, Anheim M, Bauer P, Bonnot O, et al. Recommendations for the detection and diagnosis of Niemann-Pick disease type C: An update. Neurol Clin Pract 2017; 7: 499–511.

Patterson MC, Vecchio D, Prady H, Abel L, Wraith JE. Miglustat for treatment of Niemann-Pick C disease: a randomised controlled study. Lancet Neurol 2007; 6: 765–772.

Pineda M, Perez-Poyato MS, O’Callaghan M, Vilaseca MA, Pocovi M, Domingo R, et al. Clinical experience with miglustat therapy in pediatric patients with Niemann-Pick disease type C: A case series. Mol Genet Metab 2010; 99: 358–366.

Rottach KG, von Maydell RD, Das VE, Zivotofsky AZ, Discenna AO, Gordon JL, et al. Evidence for independent feedback control of horizontal and vertical saccades from Niemann-Pick type C disease. Vision Res 1997; 37: 3627–3638.

Rucker JC, Shapiro BE, Han YH, Kumar AN, Garbutt S, Keller EL, et al. Neuro-ophthalmology of late-onset Tay-Sachs disease (LOTS). Neurology 2004; 63: 1918–1926.

Rucker JC, Ying SH, Moore W, Optican LM, Büttner-Ennever J, Keller EL, et al. Do brainstem omnipause neurons terminate saccades? Ann N Y Acad Sci 2011; 1233: 48–57.

Salsano E, Umeh C, Rufa A, Pareyson D, Zee DS. Vertical supranuclear gaze palsy in Niemann-Pick type C disease. Neurol Sci Off J Ital Neurol Soc Ital Soc Clin Neurophysiol 2012; 33: 1225–1232.

Schillevoort I, de Boer A, Herings RM, Roos RA, Jansen PA, Leufkens HG. Antipsychotic-induced extrapyramidal syndromes. Risperidone compared with low-and high-potency conventional antipsychotic drugs. Eur J Clin Pharmacol 2001; 57: 327–331.

Schillevoort I, de Boer A, Herings RM, Roos RA, Jansen PA, Leufkens HG. Risk of extrapyramidal syndromes with haloperidol, risperidone, or olanzapine. Ann Pharmacother 2001; 35: 1517–1522.

Schmitz-Hübsch T, Giunti P, Stephenson DA, Globas C, Baliko L, Saccà F, et al. SCA Functional Index: a useful compound performance measure for spinocerebellar ataxia. Neurology 2008; 71: 486–492.

Schneider E, Villgrattner T, Vockeroth J, Bartl K, Kohlbecher S, Bardins S, et al. EyeSeeCam: An Eye Movement-Driven Head Camera for the Examination of Natural Visual Exploration. Annals of the New York Academy of Sciences 2009; 1164: 461–467.

Sidhu R, Kell P, Dietzen DJ, Farhat NY, Do AND, Porter FD, et al. Application of N-palmitoyl-O-phosphocholineserine for diagnosis and assessment of response to treatment in Niemann-Pick type C disease. Mol Genet Metab 2020; 129: 292–302.

Solomon D, Winkelman AC, Zee DS, Gray L, Büttner-Ennever J. Niemann-Pick type C disease in two affected sisters: ocular motor recordings and brain-stem neuropathology. Ann N Y Acad Sci 2005; 1039: 436–445.

Subramony SH. SARA--a new clinical scale for the assessment and rating of ataxia. Nat Clin Pract Neurol 2007; 3: 136–137.

Taghdiri F, Chung J, Irwin S, Multani N, Tarazi A, Ebraheem A, et al. Decreased Number of Self-Paced Saccades in Post-Concussion Syndrome Associated with Higher Symptom Burden and Reduced White Matter Integrity. J Neurotrauma 2018; 35: 719–729.

Van Gisbergen JA, Robinson DA, Gielen S. A quantitative analysis of generation of saccadic eye movements by burst neurons. J Neurophysiol 1981; 45: 417–442.

Vanier MT, Millat G. Niemann-Pick disease type C. Clin. Genet. 2003; 64: 269–81.

Vanier MT. Niemann-Pick disease type C. Orphanet J Rare Dis 2010 3; 5:16.

Vanier MT. Complex lipid trafficking in Niemann-Pick disease type C. J Inherit Metab Dis 2015; 38: 187–199.

Weyer A, Abele M, Schmitz-Hübsch T, Schoch B, Frings M, Timmann D, et al. Reliability and validity of the scale for the assessment and rating of ataxia: a study in 64 ataxia patients. Mov. Disord. 2007; 22: 1633–1637.

Xiong H, Higaki K, Wei C-J, Bao X-H, Zhang Y-H, Fu N, et al. Genotype/phenotype of 6 Chinese cases with Niemann-Pick disease type C. Gene 2012; 498: 332–335.

